# Etiological profile of digital necrosis in internal medicine: a retrospective study of 44 cases

**DOI:** 10.64898/2026.11.21.26368639

**Authors:** Sana Toujani, Youssef Belhaj Hassen, Amira El Ouni, Youssef Hnen, Cherifa Abdelkefi, Asma Tekaya, Thara Larbi, Zeineb Meddeb, Salwa Hamzaoui

## Abstract

**Background:** To describe the etiological profile of digital necrosis in an internal medicine setting and to assess the respective diagnostic yield of the clinical and paraclinical components of the etiological work-up.

**Methods:** A descriptive, retrospective, single-centre study of 44 patients admitted for one or more digital necroses to an internal medicine department between 1997 and 2022. For each patient, clinical data and documented complementary investigations were reviewed; an item was considered contributory when its positivity oriented the etiological diagnosis.

**Results:** The 44 patients comprised 23 women and 21 men (male-to-female ratio 0.91), with a mean age of 51 ± 14 years. Raynaud’s phenomenon preceded the necrosis in 36.4% of cases. The necrosis involved at least two fingers (72.7%), the index finger being most frequently affected (56.8%); associated toe necrosis was present in 18.1% of cases. Connective tissue diseases were the leading cause (29.5%), dominated by systemic sclerosis, followed by thromboangiitis obliterans (18.1%), malignancy (15.9%) and vasculitis (9.1%). The necrosis revealed the underlying disease in 35 patients (79.5%), and the work-up remained inconclusive in 11.3% of cases. History taking and physical examination were contributory in 61.4% and 43.2% of cases, ahead of biological (37.1%), morphological (40%) and histopathological (36.4%) investigations.

**Conclusions:** The etiological profile of digital necrosis in internal medicine is dominated by connective tissue diseases, the necrosis frequently revealing the underlying disorder. History taking and physical examination provide the highest diagnostic yield and should guide a targeted prescription of investigations.

## Introduction

Digital necrosis reflects downstream vascular involvement, either macroangiopathic or microangiopathic. Its positive diagnosis is usually straightforward and clinical: the ischemic nature of a digital lesion must be recognized, sometimes preceded by paresthesia, pain, or Raynaud’s phenomenon suggesting a vascular origin [1,2]. Rarer than toe necrosis, it constitutes the presenting symptom of a wide range of systemic or locoregional conditions that it may reveal or complicate.

In the absence of a consensus codifying the diagnostic approach, and given the heterogeneity of the etiological spectrum, the search for the cause remains a genuine challenge, particularly when the necrosis is the presenting manifestation [3]. We propose to study these etiologies in a retrospective series of 44 cases, and to assess, which has not previously been reported, the respective diagnostic yield of the different components of the etiological work-up.

## Patients and methods

This was a descriptive, retrospective study conducted in the department of internal medicine of the Mongi Slim University Hospital of La Marsa (Tunisia) between 1997 and 2022. Patients hospitalized for investigation of one or more digital necroses, with or without toe necrosis, as well as those who developed necrosis during follow-up of a known condition, were included.

All patients underwent a complete clinical examination, a complete blood count, liver function tests, and a chest radiograph; erythrocyte sedimentation rate and C-reactive protein were measured in nearly all of them (90.9%). The other investigations were performed selectively, according to the clinical context and the results of first-line tests, consistent with the retrospective nature of the study: laboratory investigations (immunological work-up, cryoglobulinemia testing, viral serologies, specialized assays), morphological investigations (ultrasonography, computed tomography, capillaroscopy), and histopathological investigations.

The initial French-to-English translation of this manuscript was performed with the assistance of an AI language model (Claude, Anthropic); all translated content was subsequently reviewed, verified, and approved by the authors for scientific and factual accuracy, and the AI model was not used for study design, data analysis, or interpretation of results.

## Results

The 44 patients comprised 23 women and 21 men, giving a male-to-female ratio of 0.91, with a mean age of 51 ± 14 years (range 14–74). Seven patients (15.9%) had an occupation exposing them to microtrauma or vibration (heavy-goods vehicle driver, seamstress, mechanic, construction worker, welder, musician). The mean time to consultation was 12 ± 8 days, and the mean time to etiological diagnosis was 19 ± 13 days.

Twenty-one patients (47.7%) were smokers, with a mean consumption of 39 ± 21 pack-years (range 10– 80); hookah use and neffa (oral snuff) use were each reported by one patient. Two patients reported regular cannabis use, and one patient reported use of cocaine, heroin, and buprenorphine. A history of self-inflicted skin cutting was found in two patients, and of tattooing in one patient. Raynaud’s phenomenon preexisted in 16 patients (36.4%), with a mean duration of 42 ± 24 months. The necrosis involved at least two fingers in 32 patients (72.7%), the index finger being the most frequently affected (n = 25, 56.8%); associated toe necrosis was present in eight cases (18.1%). The distribution of etiologies is presented in **Table 1**.

**Table 1.** Distribution of the etiologies of digital necrosis (n = 44).

|  |  |  |
|---|---|---|
| Connective tissue diseases | 13 | 29,5 |
| Systemic sclerosis | 8 | 18,1 |
| Sharp syndrome | 3 | 6,8 |
| Systemic lupus erythematosus | 1 | 2,3 |
| Rheumatoid arthritis | 1 | 2,3 |
| Thromboangiitis obliterans | 8 | 18,1 |
| Malignancies | 7 | 15,9 |
| Primary vasculitis | 4 | 9,1 |
| Drug-induced cause (norepinephrine) | 2 | 4,5 |
| Toxic cause (buprenorphine) | 1 | 2,3 |
| Embolic cardiopathy | 1 | 2,3 |
| Miscellaneous causes* | 3 | 6,8 |
| Undetermined | 5 | 11,3 |

The connective tissue diseases were the leading cause (13 cases, 29.5%): systemic sclerosis (n = 8, including one case of RACAND syndrome), Sharp syndrome (n = 3), systemic lupus erythematosus (n = 1), and rheumatoid arthritis complicated by rheumatoid vasculitis (n = 1). Next were thromboangiitis obliterans (n = 8, 18.1%, including one cannabis user), active malignancies (n = 7, 15.9%: lung n = 2, breast, ovary, melanoma, hepatocellular carcinoma, and cholangiocarcinoma), and primary vasculitis (n = 4, 9%: giant cell arteritis n = 2, cryoglobulinemic vasculitis n = 2). A drug-induced cause was identified in two patients exposed to norepinephrine (4.5%), a toxic cause in one patient who had self-injected buprenorphine (2.3%), and an embolic cardiopathy (infective endocarditis) in one patient (2.3%). Miscellaneous causes included pheochromocytoma, antiphospholipid syndrome, and arteriosclerosis (one case each). Finally, the work-up remained inconclusive in five patients (11.3%). Digital necrosis was the presenting and revealing manifestation of the etiology in 35 patients (79.5%); nine patients (20.5%) were already being followed for the causal disease.

In terms of treatment, symptomatic management relied mainly on analgesics (38 patients, 86.4%), with paracetamol being the most commonly used, followed by step II and step III analgesics, a combination of steps sometimes being necessary (Figure 1). Next came antiplatelet therapy and calcium channel blockers (35 patients each, 79.5%), curative heparin therapy (30 patients, 68.2%), and iloprost (13 patients, 29.5%). Etiological treatment was instituted according to the cause: corticosteroid therapy (14 patients, 31.8%), immunosuppressants, chemotherapy, radiotherapy, or appropriate cancer surgery. The frequency of use of the different treatments is detailed in Figure 2. Digital amputation was required in 16 patients (36.4%), and nine patients (20.5%) died during follow-up.

**Figure 1.**
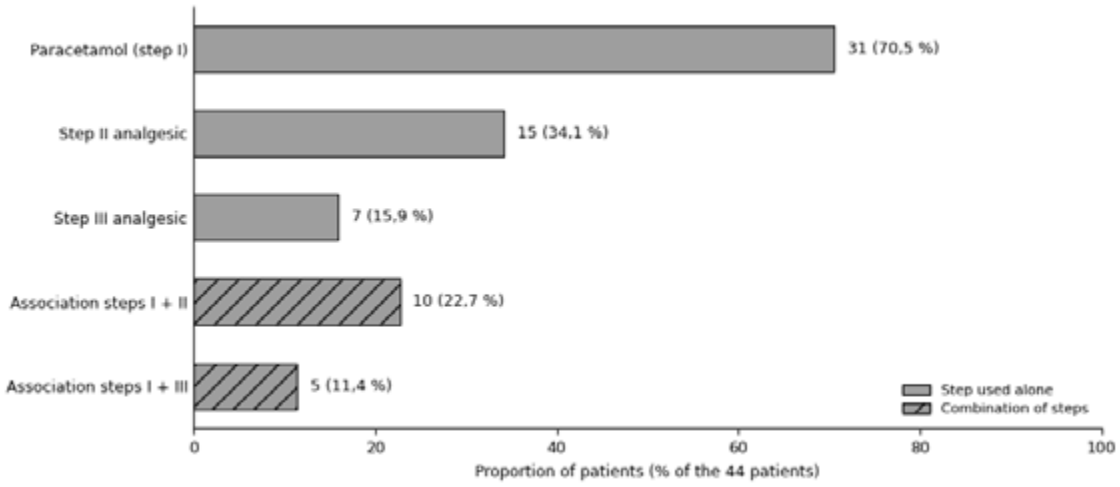
Distribution of the analgesic steps used in treated patients (n = 44).

**Figure 2.**
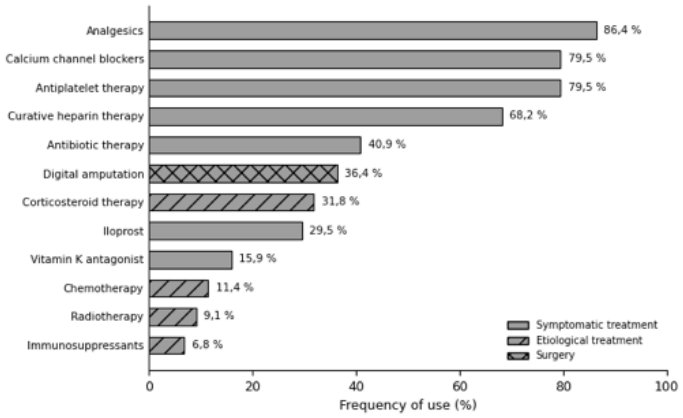
Therapeutic modalities used in the management of digital necrosis (n = 44).

On the diagnostic level (**Table 2**), history taking including age, habits, medical history, and functional symptoms was contributory in 27 patients (61.4%), and physical examination in 19 patients (43.2%), with skin examination alone being contributory in 27.3% of cases. Among laboratory investigations (overall contribution 37.1%), the immunological work-up, requested in 70.5% of cases, was contributory in 29% of cases; blood culture, measurement of methoxylated derivatives, and viral serologies allowed identification of infective endocarditis, pheochromocytoma, and cryoglobulinemic vasculitis, respectively. Morphological investigations were contributory in 40% of cases, with capillaroscopy showing the best targeted yield (42.9% for only 15.9% performance). Histopathological investigations were contributory in 36.4% of cases, with temporal artery biopsy confirming giant cell arteritis in both cases in which it was performed, and lymph node biopsy revealing metastasis from a poorly differentiated carcinoma and a melanoma.

**Table 2.**
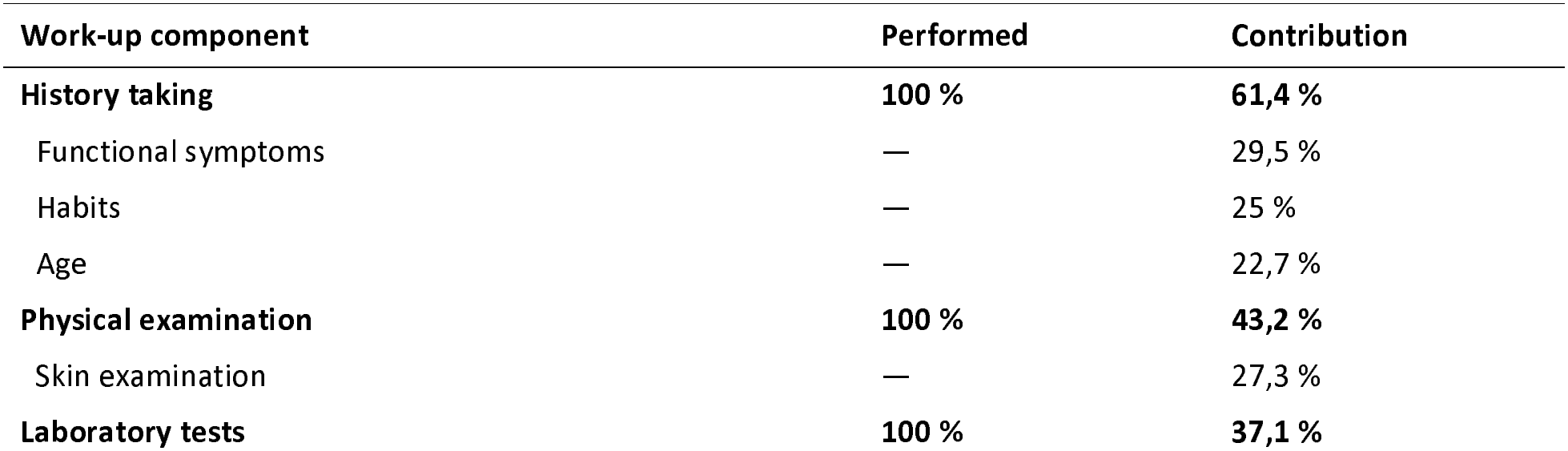
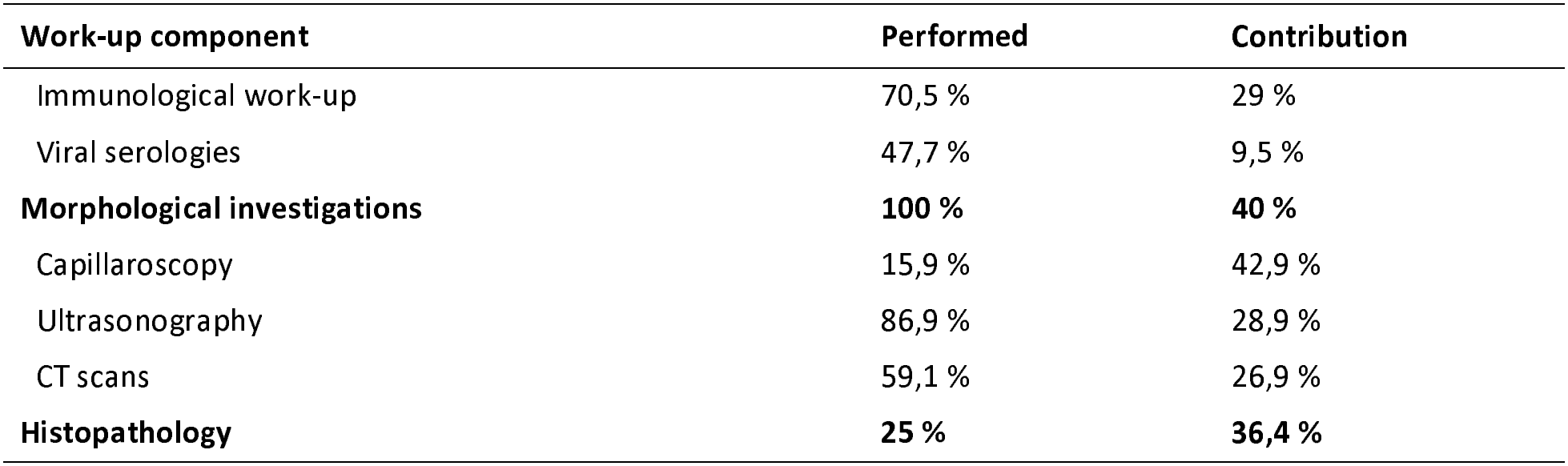
Frequency of performance and degree of diagnostic contribution of the main components of the etiological work-up.

| Work-up component | Performed | Contribution |
|---|---|---|
| **History taking** | **100 %** | **61,4 %** |
| Functional symptoms | — | 29,5 % |
| Habits | — | 25 % |
| Age | — | 22,7 % |
| **Physical examination** | **100 %** | **43,2 %** |
| Skin examination | — | 27,3 % |
| **Laboratory tests** | **100 %** | **37,1 %** |
| Immunological work-up | 70,5 % | 29 % |
| Viral serologies | 47,7 % | 9,5 % |
| **Morphological investigations** | **100 %** | **40 %** |
| Capillaroscopy | 15,9 % | 42,9 % |
| Ultrasonography | 86,9 % | 28,9 % |
| CT scans | 59,1 % | 26,9 % |
| **Histopathology** | **25 %** | **36,4 %** |

## Discussion

This single-centre retrospective study of 44 digital necroses collected in an internal medicine department illustrates the diversity of the etiological spectrum of this manifestation and clarifies the respective contribution of clinical and paraclinical elements to the diagnostic work-up. Connective tissue diseases were the leading cause (29.5%), ahead of thromboangiitis obliterans (18.1%), malignancy (15.9%), and vasculitis (9.1%), and the necrosis revealed the underlying disease in 79.5% of cases.

The etiological distribution differs from that of several series (**Table 3**), and these discrepancies are largely explained by patient recruitment and inclusion criteria. The frequency of connective tissue diseases thus ranges from 6.5% to 71.4% across studies [4–11]: it is highest in series enriched for scleroderma [9,10] and lowest when scleroderma-related necroses are excluded, as in Cailleux et al. (20%) [6].

**Table 3.** Frequency of the main etiologies in our series and in the literature.

| **Series (n)** | **Connective tissue diseases** | **Buerger** | **Vasculitis** | **Malignancies** |
|---|---|---|---|---|
| Vayssairat [5] (86) | 40,7 % | 4,7 % | 8,1 % | 3,5 % |
| Cailleux [6] (45) | 20 % | 13,3 % | 4,4 % | 2,2 % |
| Abdallah [4] (22) | 34,8 % | 22,7 % | 22,7 % | 9,1 % |
| Staszewski [7] (92) | 6,5 % | 39,2 % | 5,4 % | 3,3 % |
| Le Besnerais [8] (100) | 10 % | 22 % | 5 % | 15 % |
| Toujani [9] (28) | 71,4 % | 10,7 % | 7,1 % | 3,6 % |
| Aydi [10] (23) | 69,5 % | 10 % | 4,3 % | 4,3 % |
| Raimbeau [11] (323) | 19,2 % | 9,3 % | 3,1 % | 6,2 % |
| **Our series (44)** | **29,5 %** | **18,1 %** | **9,1 %** | **15,9 %** |

Our intermediate figure (29.5%, including 18.1% systemic sclerosis) corresponds to an internal medicine recruitment setting in which connective tissue diseases are overrepresented. The most marked divergence concerns the cohort of Raimbeau et al. (323 patients), in which embolic cardiopathy was the leading cause (18.3%) and iatrogenic causes reached 9.6%, compared with 2.3% and 6.8%, respectively, in our series [11]. This difference is attributable to the definition used by these authors, who included hand ischemia rather than digital necrosis alone, broadening the recruitment of cardio-embolic causes and to a systematic search for iatrogenic causes, which was less thoroughly explored in earlier studies; in addition, a period effect may be involved, with the use of endovascular procedures and vasoconstrictive drugs having increased in recent years.

The frequency of thromboangiitis obliterans (18.1%) is consistent with the literature average (≈15%) and occurred in a population with heavy tobacco exposure. Malignancy, in contrast, was more frequent (15.9%) than the reported average (≈6.4%), close to the 15% reported by Le Besnerais et al. [8]; as in that cohort, bronchopulmonary cancer predominated, and most patients (57.1%) were at an advanced stage, supporting the concept of paraneoplastic acral vascular syndrome, in which age over 50 years and thrombocytosis suggest an occult malignancy [8,21]. To this framework must now be added iatrogenic acral ischemia induced by anticancer treatments themselves, particularly immune checkpoint inhibitors [21], an entity rarely described at the time of the earlier series. Finally, the proportion of cases that remained undetermined (11.3%) is comparable to the 1.6% to 22% reported in the literature [11].

These etiologies involve distinct mechanisms. In systemic sclerosis, necrosis results from early obliterative microangiopathy combining endothelial dysfunction, intimal proliferation, and intraluminal thrombosis, with young age, the diffuse cutaneous form, and anti-Scl70 antibodies favoring ulceration [14,15]. Thromboangiitis obliterans corresponds to segmental inflammatory thrombosis of small and medium-sized vessels, closely linked to tobacco use, for which smoking cessation remains the key treatment [16]; chronic cannabis use may contribute to it or induce arteritis specific to young individuals [17,23]. In paraneoplastic forms, tumor-related hypercoagulability, immune complex vasculitis, and secretion of vasoconstrictive substances have been implicated [8,21], while α-adrenergic vasoconstriction accounts for the necroses associated with norepinephrine [18].

In terms of treatment, our management approach, based primarily on calcium channel blockers and intravenous iloprost, is consistent with the 2023 EULAR recommendations, which define a vasoactive continuum from calcium channel blockers to phosphodiesterase type 5 inhibitors and prostacyclin analogues for active ulcers, with endothelin receptor antagonists reducing the occurrence of new ulcers in scleroderma [20,24,25].

Our study has several limitations. Its retrospective design, spanning a 25-year period, exposes it to missing data and heterogeneity in the technical platform, partially offset by the exclusion of unusable records and re-evaluation according to current classification criteria. The single-centre design and limited sample size restrict the generalizability of the results, and the definition of diagnostic contribution remains partly operator-dependent. The low proportion of cardio-embolic and iatrogenic causes probably reflects a recruitment bias, as the diagnosis of these conditions is often made upstream of an internal medicine department, similar to the underrepresentation of occupational causes observed by other teams [11].

Given the respective contribution of the investigations in our series, a hierarchical approach can be proposed. History taking and physical examination, contributory in 61.4% and 43.2% of cases, should form the foundation, with skin examination alone providing orientation in 27.3% of cases. In the absence of orientation, cardiac investigation (electrocardiogram, transthoracic followed by transesophageal echocardiography) allows an embolic cardiopathy to be ruled out. Second-line investigations should be guided by the clinical context: immunological work-up and capillaroscopy in young women, in whom the targeted yield is high [19,22], and screening for occult malignancy in older patients. Conversely, thrombophilia work-up and homocysteine measurement, which were consistently negative, do not appear to be justified as first-line investigations [11]. Such a sequence, to be validated by prospective multicentre studies, would help rationalize the etiological work-up of digital necrosis.

## Conclusions

Digital necrosis is a rare but severe manifestation, with an easy positive diagnosis but a complex etiological work-up. In internal medicine, its profile is dominated by connective tissue diseases, followed by thromboangiitis obliterans, malignancy, and vasculitis, with the necrosis revealing the underlying disease in nearly four out of five cases. History taking and physical examination provide the best diagnostic yield and should guide a rational prescription of investigations in order to initiate appropriate treatment early and preserve functional prognosis.

## Data Availability

All data produced in the present study are available upon reasonable request to the authors

## Declarations

### Ethics approval and consent to participate

The study protocol was approved by the Ethics Committee of Mongi Slim Hospital, La Marsa, Tunis, Tunisia. Given the retrospective, non-interventional design of the study, based on anonymised data extracted from medical records, the requirement for individual written informed consent was waived by the same committee. All procedures were conducted in accordance with the Declaration of Helsinki (1964) and its later amendments. This study did not involve animals.

### Data availability

The data supporting the findings of this study are available from the authors upon reasonable request.

## List of abbreviations

CT: Computed tomography
EULAR: European Alliance of Associations for Rheumatology (formerly the European League Against Rheumatism)

